# Electrophysiological Features and Catheter Ablation for Supraventricular Tachyarrhythmias in Patients with Fontan Circulation: A Multicenter Study

**DOI:** 10.64898/2026.03.23.26349127

**Authors:** Jae-Sun Uhm, Mi Kyoung Song, Ji-Eun Ban, Seung Min Baek, Taehyun Hwang, Seunghoon Cho, Hanjin Park, Daehoon Kim, Hee Tae Yu, Tae-Hoon Kim, Boyoung Joung, Hui-Nam Pak, Nuri Tchah, Na Hyun Lee, Chang Sin Kim, Su-Jin Park, Jo Won Jung, Jae Young Choi, Eun Jung Bae

**Affiliations:** Department of Cardiology, Severance Hospital, Yonsei University College of Medicine, Seoul, Korea; Department of Pediatrics, Seoul National University Children’s Hospital, Seoul National University College of Medicine, Seoul, Korea; Department of Pediatrics, Sejong General Hospital, Bucheon, Gyeonggi-do, Korea; Division of Pediatric Cardiology, Severance Hospital, Yonsei University College of Medicine, Seoul, Korea

**Author notes:** Jae-Sun Uhm and Mi Kyoung Song contributed equally to this work as the first author. **Corresponding author:** Jae-Sun Uhm, MD, PhD, Department of Cardiology, Severance Hospital, Yonsei University College of Medicine, 50-1 Yonsei-ro Seodaemun-gu, Seoul, 03722, Republic of Korea; Eun Jung Bae, MD, PhD, Department of Pediatrics, Seoul National University Children’s Hospital, Seoul National University College of Medicine, 101 Daehak-ro Jongno-gu, Seoul, 03080, Republic of Korea.

**Keywords:** arrhythmia, atrial tachycardia, catheter ablation, Fontan procedure, intra-atrial reentrant tachycardia, single ventricle, supraventricular tachycardia

## Abstract

**Background:** Patients with Fontan circulation experience significant morbidity from supraventricular tachyarrhythmias (SVTs). However, the electrophysiological features of SVT and the efficacy and safety of catheter ablation in patients with Fontan circulation are poorly understood. This study aimed to elucidate the electrophysiological features of SVT and evaluate the efficacy and safety of catheter ablation in patients with Fontan circulation.

**Methods:** Forty-nine patients (age, 29.2±10.0 years; 27 males) with functional single ventricle and Fontan circulation who had undergone electrophysiological study for SVT were retrospectively enrolled. Parameters analyzed included underlying congenital heart disease, Fontan type, conduit puncture technique, tachycardia mechanisms, tachycardia origin site, acute success rate, procedure-related complications, and recurrence.

**Results:** Fifty-nine SVTs were induced, and 69 catheter ablations were performed. The Fontan types included atriopulmonary connection (APC, 18.4%), lateral tunnel (LT, 38.8%), and extracardiac conduit (ECC, 42.9%). Inducible tachycardias included intra-atrial reentrant tachycardia (IART, 39.0%), focal atrial tachycardia (AT, 28.8%), atrioventricular reentrant tachycardia (11.9%), atrioventricular nodal reentrant tachycardia (10.2%), and atrioventricular reciprocating tachycardia involving the twin atrioventricular nodes (10.2%). The right atrial (RA) lateral wall was the most common location of IART and focal AT. The acute success and complication rates were 73.5% and 4.1%, respectively. Recurrence rate was 34.7% during follow-up of 78.0±71.9 months. The cumulative recurrence rate was significantly lower in patients who underwent LT or ECC Fontan procedures than in those who underwent the APC Fontan procedure (*P*<0.001).

**Conclusions:** Catheter ablation for SVT is effective and safe in patients who have undergone LT and ECC Fontan procedures.

**What is known?:**

- The incidence of arrhythmia is higher in patients with Fontan circulation than the general population.
- Catheter ablation for arrhythmia in patients undergoing lateral tunnel or extracardiac conduit Fontan procedures is more challenging because of the complex cardiovascular anatomy and limited accessibility.

**What the study adds:**

- Intra-atrial reentrant tachycardia (IART) and focal atrial tachycardia (AT) were the most common supraventricular tachycardias (SVT) in patients with Fontan circulation.
- The right atrial lateral wall was the most common site of critical IART isthmus and focal AT origin in patients with Fontan circulation.
- Catheter ablation for SVT was effective and safe in patients who underwent the lateral tunnel or extracardiac conduit Fontan procedures.

## Introduction

The incidence of arrhythmia is higher in adult patients with complex congenital heart disease than the general population.^1,2^ Advances in surgical and interventional techniques have improved the survival rate of patients with complex congenital heart. Consequently, arrhythmias have become a significant concern. After the first Fontan procedure for a functional single ventricle (FSV) was performed in 1968,^3^ surgical techniques have evolved from atriopulmonary connection (APC) to lateral tunnel (LT) and extracardiac conduit (ECC), reducing arrhythmogenicity.^4,5^ However, these modifications have rendered atrial access technically challenging for invasive procedures, making catheter ablation for arrhythmia in patients undergoing LT or ECC Fontan procedures more demanding because of the complex cardiovascular anatomy and limited accessibility. Despite the growing population of adults with FSV and Fontan circulation and the high burden of supraventricular tachyarrhythmia (SVT), data regarding SVT electrophysiological characteristics and the efficacy and safety of catheter ablation in these patients remain limited. This study aimed to elucidate the electrophysiological features of SVT and the efficacy and safety of catheter ablation in patients with FSV and Fontan circulation.

## Methods

This retrospective multicenter cohort study was approved by the Institutional Review Board of Severance Hospital, Seoul National University Children’s Hospital, and Sejong General Hospital (IRB number: 4-2025-1595, 2602-118-1718, 2026-02-006) and conformed to the principles outlined in the Declaration of Helsinki. All patients provided written informed consent to undergo an electrophysiological study and catheter ablation for SVT.

Patients with FSV and Fontan circulation (APC, LT, and ECC) who had undergone electrophysiological study for SVT from July 2011 to November 2025 were retrospectively enrolled from three tertiary hospitals. SVT includes intra-atrial reentrant tachycardia (IART), focal atrial tachycardia (AT), atrioventricular (AV) nodal reentrant tachycardia (AVNRT), AV reentrant tachycardia (AVRT), and AV reciprocating tachycardia involving twin AV nodes (AVRT-TN). Patients who underwent catheter ablation for atrial fibrillation or ventricular tachycardia were excluded. Demographic and clinical data were collected from the medical records.

Before the procedure, transthoracic echocardiography and computed tomography (CT) were performed. The procedures were performed under deep sedation or general anesthesia. Electrophysiological catheters were placed at the discretion of the operator. A decapolar or quadripolar catheter was placed in the left or right pulmonary artery adjacent to the atrium through the femoral or jugular vein and Fontan pathway for atrial sensing and reference. When adequate atrial signal could not be obtained from the pulmonary artery, a quadripolar catheter was placed in the esophagus adjacent to the atrium for atrial sensing and reference. A quadripolar catheter was placed in the ventricle via the femoral artery and aorta if ventricular sensing and pacing were required. A quadripolar catheter was placed within the Fontan pathway through the femoral vein to facilitate atrial sensing and pacing. In instances where atrial sensing and pacing were impossible through catheter placement, a quadripolar catheter was placed in the esophagus adjacent to the atrium. Three-dimensional mapping was performed using the CARTO system (Johnson & Johnson Med Tech, Diamond Bar, California, USA) or the EnSite system (Abbott, Minneapolis, Minnesota, USA). Baseline electrophysiological studies were conducted, and tachycardia induction was performed using programmed electrical stimuli with or without isoproterenol infusion. In patients with inducible tachycardia who have undergone LT or ECC Fontan procedures, the Fontan conduit was punctured under intracardiac or transesophageal echocardiography guidance, as outlined below (Figure 1); (1) Fontan conduit angiography (Figure 2 A); (2) Fontan puncture using the Swartz sheath and Brockenbrough needle; (3) in cases of failed puncture, a modified approach was adopted using the Swartz sheath, Brockenbrough needle, and a cut stylet (stylet cut obliquely 1–2 mm from the Brockenbrough needle tip; Figure 2 B and C); (4) if puncture was successful but inadequate for Swartz sheath introduction, staged balloon dilation of the puncture site was performed (Figure 2 D).^6,7^ In patients with cavoatrial overlap, transcaval puncture was performed through a segment of the inferior vena cava adjacent to the atrial wall immediately below the inferior ECC margin. After accessing the atrium, the activated clotting time was maintained between 300 and 350 s using heparin. Three-dimensional activation mapping was performed during tachycardia using a multi-electrode mapping catheter. In cases of reentrant tachycardia, catheter ablation was performed if reentry circuits and their critical isthmus were identified. In cases of focal AT, catheter ablation was performed if focal AT origins were identified. The procedural endpoint was non-inducibility and/or tachycardia termination during catheter ablation. The patients were followed-up at the outpatient clinic at 3- or 6-month intervals. Electrograms (ECGs) were recorded at every visit. Holter or patch ECG monitoring was performed when patients reported of symptoms such as palpitations, chest discomfort, dizziness, or syncope.

**Figure 1.**
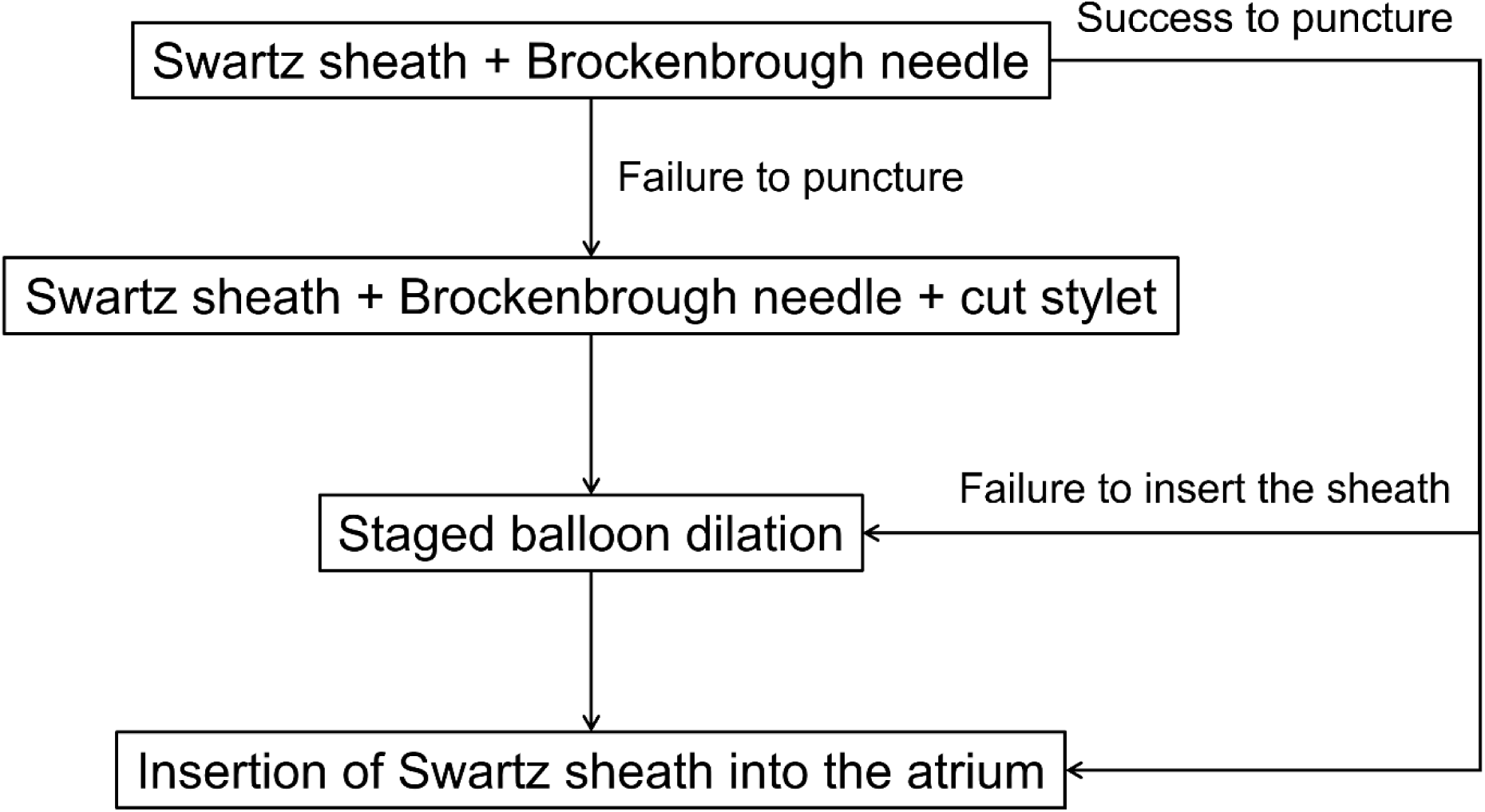
Workflow for Fontan conduit puncture.

**Figure 2.**
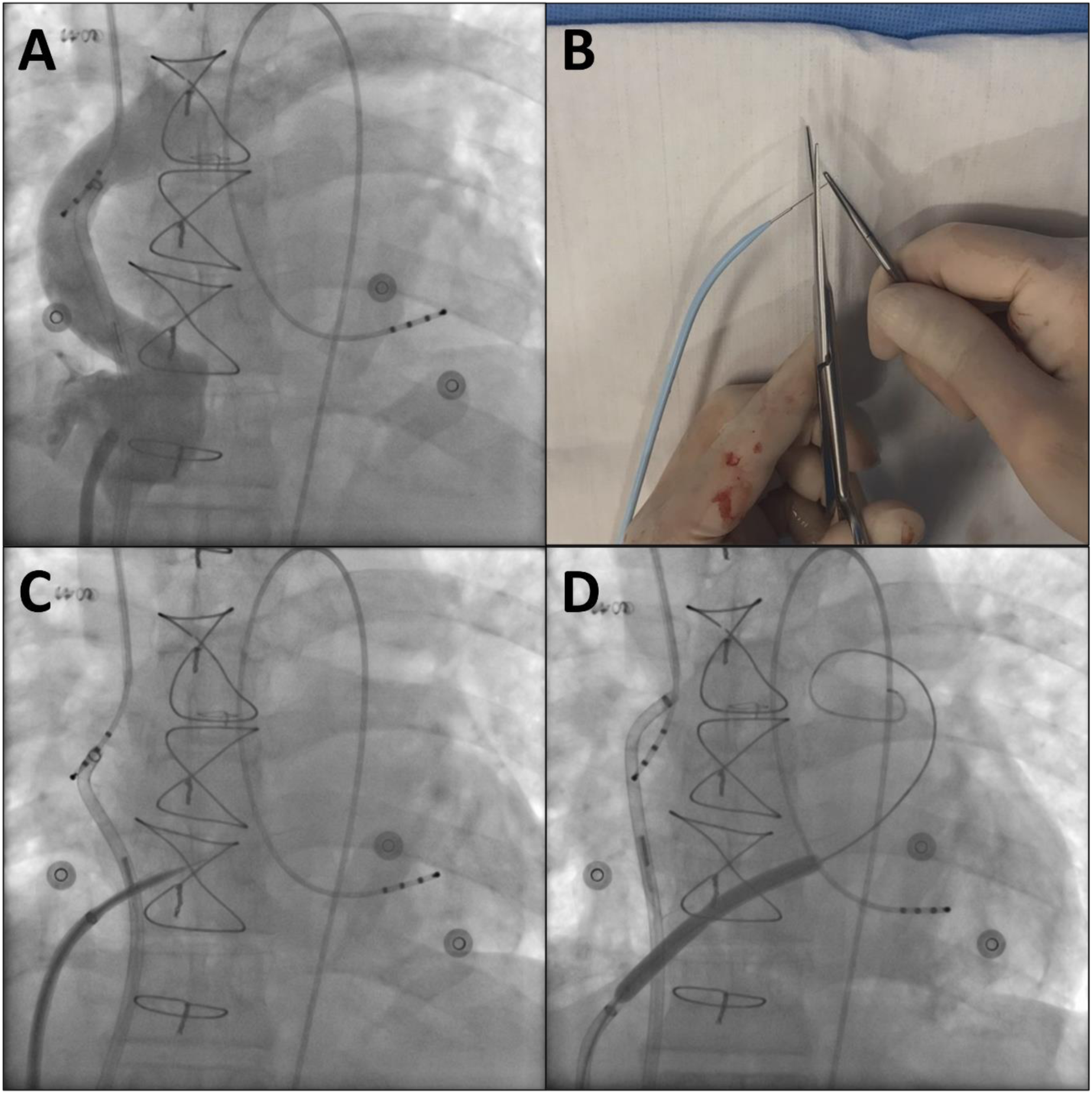
Process of extracardiac conduit puncture. **A.** The Fontan conduit angiogram. **B.** A representative image of the stylet cut. **C.** An image showing the Fontan conduit puncture using Swartz sheath, Brockenbrough needle, and cut stylet under intracardiac echocardiography guidance. **D.** An image showing the balloon dilation.

The characteristics of the underlying congenital heart disease, Fontan conduit puncture techniques, tachycardia mechanisms, and locations of critical isthmus in reentrant tachycardia or focal AT origins were analyzed. Acute success, complications, and SVT recurrence rates were analyzed according to arrhythmia and Fontan type. Cumulative SVT recurrence was compared across Fontan types.

### Statistical Analysis

Continuous variables are expressed as mean ± standard deviation and compared using the Kruskall–Wallis test. Categorical variables are expressed as numbers (percentages) and compared using Fisher’s exact test. The Kaplan–Meier method was used to compare cumulative SVT recurrence. Statistical significance was set at *P*<0.05. Data were analyzed using the Statistical Package for the Social Sciences, version 27.0 (IBM Corp., Armonk, New York, USA).

## Results

This study included 49 patients (mean age, 29.2±10.0 years; 27 males). Fifty-nine SVTs were induced, and 69 catheter ablation procedures were performed in these patients. Table 1 presents the baseline characteristics of the patients according to Fontan type. Fontan types were as follows: APC (nine patients), LT (19 patients), and ECC (21 patients). Patients who underwent the APC Fontan procedure were significantly older than those who underwent the LT or ECC Fontan procedure. The proportion of the men who underwent the ECC Fontan procedure were significantly more than those who underwent the APC or LT Fontan procedure. The underlying congenital heart diseases were as follows (in order of frequency): complicated AV septal defect, complicated double-outlet right ventricle, tricuspid atresia, pulmonary atresia with intact ventricular septum, mitral atresia, hypoplastic left heart syndrome, double-inlet left ventricle, Ebstein anomaly, complicated congenitally corrected transposition of the great arteries, and unrepairable “Swiss-cheese” multiple ventricular septal defects. The prevalence of sinus node dysfunction was significantly higher in patients who underwent the APC Fontan procedure than in those who underwent the LT or ECC Fontan procedure (*P*=0.010). Age at the time of the Fontan procedure, ventricular ejection fraction, and time interval from Fontan procedure to catheter ablation did not significantly differ across Fontan types. Table 2 lists the Fontan conduit puncture methods and electrophysiological features. In 8 of 9 patients who underwent the APC Fontan procedure, Fontan puncture was not required as IART originated from the right atrium (RA) or systemic venous atrium. In the remaining one patient who underwent the APC Fontan procedure, baffle puncture was performed as SVT originated from the pulmonary venous atrium. The LT baffle was successfully punctured in 15 out of 15 patients. Baffle puncture was not required in four of 19 patients who underwent the LT Fontan procedure as the focal AT originated from the Fontan pathway. ECC puncture was successful in 19 of 20 patients. For ECC puncture, the inferior vena cava approach was used in all patients except one patient in whom the superior vena cava approach was used. In one patient who underwent the ECC Fontan procedure, ECC puncture was not required as inducible AT was not sustained. For LT baffle puncture, the Brockenbrough needle only was the most effective method. For ECC puncture, “the Brockenbrough needle with cut stylet and staged balloon dilation” was the most effective method. Transcaval cardiac puncture was used in 15.0% of the patients who underwent the LT or ECC Fontan procedures. In one patient who underwent the ECC Fontan procedure, conduit puncture failed as both femoral veins were interrupted. Multiple tachycardias were induced in 22.4% of patients. Inducible tachycardias were IART, focal AT, AVRT, AVNRT, and AVRT-TN (in order of frequency). IART was more common in the patients who underwent APC than in those who underwent LT or ECC (*P*=0.007). Table 3 lists the locations of critical IART isthmuses and focal AT origins. An RA lateral wall was the most common location of IARTs (Figure 3, 4, and 5) and focal ATs (Figure 6). The RA lateral wall as an origin of IART or focal AT was significant frequent in the patients who underwent the APC or LT Fontan procedure than in those who underwent ECC Fontan procedure (*P*=0.033). Multiple IART circuits and focal ATs were identified around the surgical scar and maze ablation lines in two patients with SVT after prior Fontan conversion (APC to ECC) with maze procedure. The slow-fast and fast-slow AVNRTs were induced in six and one of the seven patients with AVNRT, respectively. Electrogram-guided catheter ablation for the slow pathway was successfully performed at the right infero- and anterosepta in five and one of the seven patients, respectively. In one patient, catheter ablation for the slow pathway was failed. The posterior AV node was ablated in five of the six patients with AVRT-TN, because the anterior AV node was the dominant AV node.

**Figure 3.**
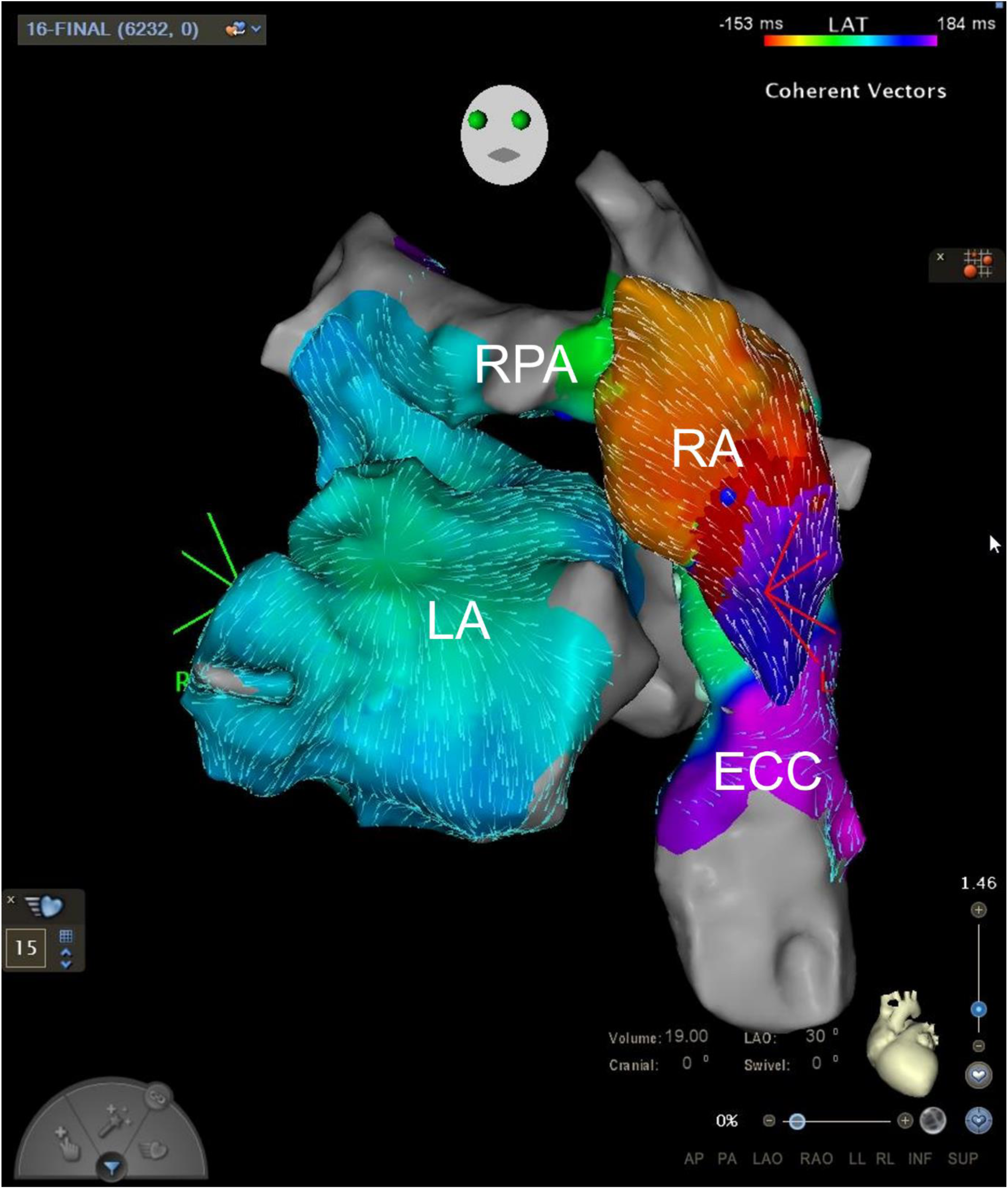
Activation map of IART in a female in her 40s who had undergone Fontan conversion operation from APC to ECC for complicated AVSD. The critical isthmus (red area) was located at the RA remnant tissue after RA resection during Fontan conversion operation. APC: atriopulmonary connection; AVSD: atrioventricular septal defect; ECC: extracardiac conduit; IART: intra-atrial reentrant tachycardia; LA: left atrium; RA: right atrium; RPA: right pulmonary artery

**Figure 4.**
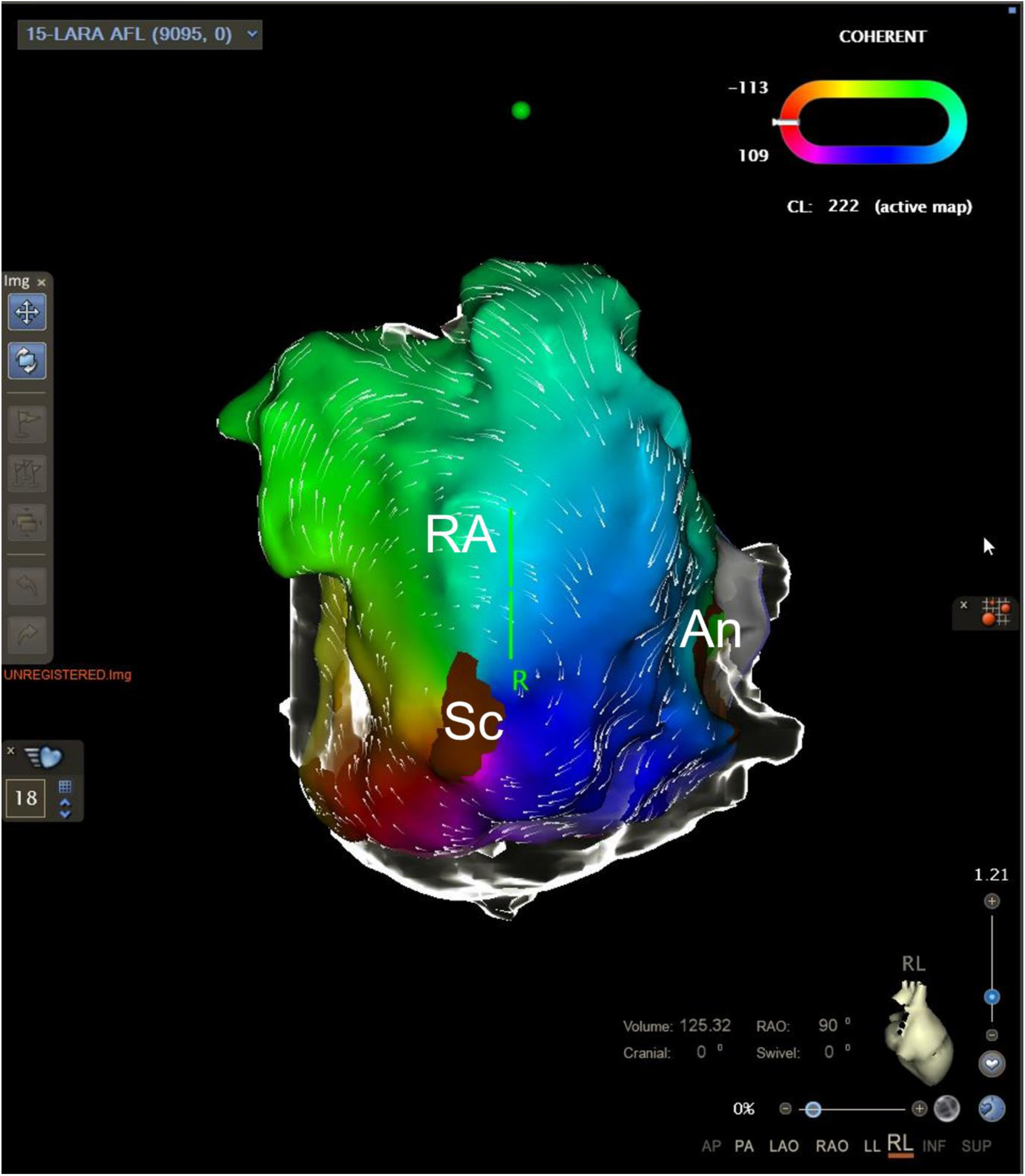
Activation map RA scar-related reentrant tachycardia in a female in her 30s with ECC Fontan for complicated AVSD. Catheter ablation for RA scar-to-inferior vena cava was performed. An: annulus of the atrioventricular valve; AVSD: atrioventricular septal defect; ECC: extracardiac conduit; RA: right atrium; Sc: scar

**Figure 5.**
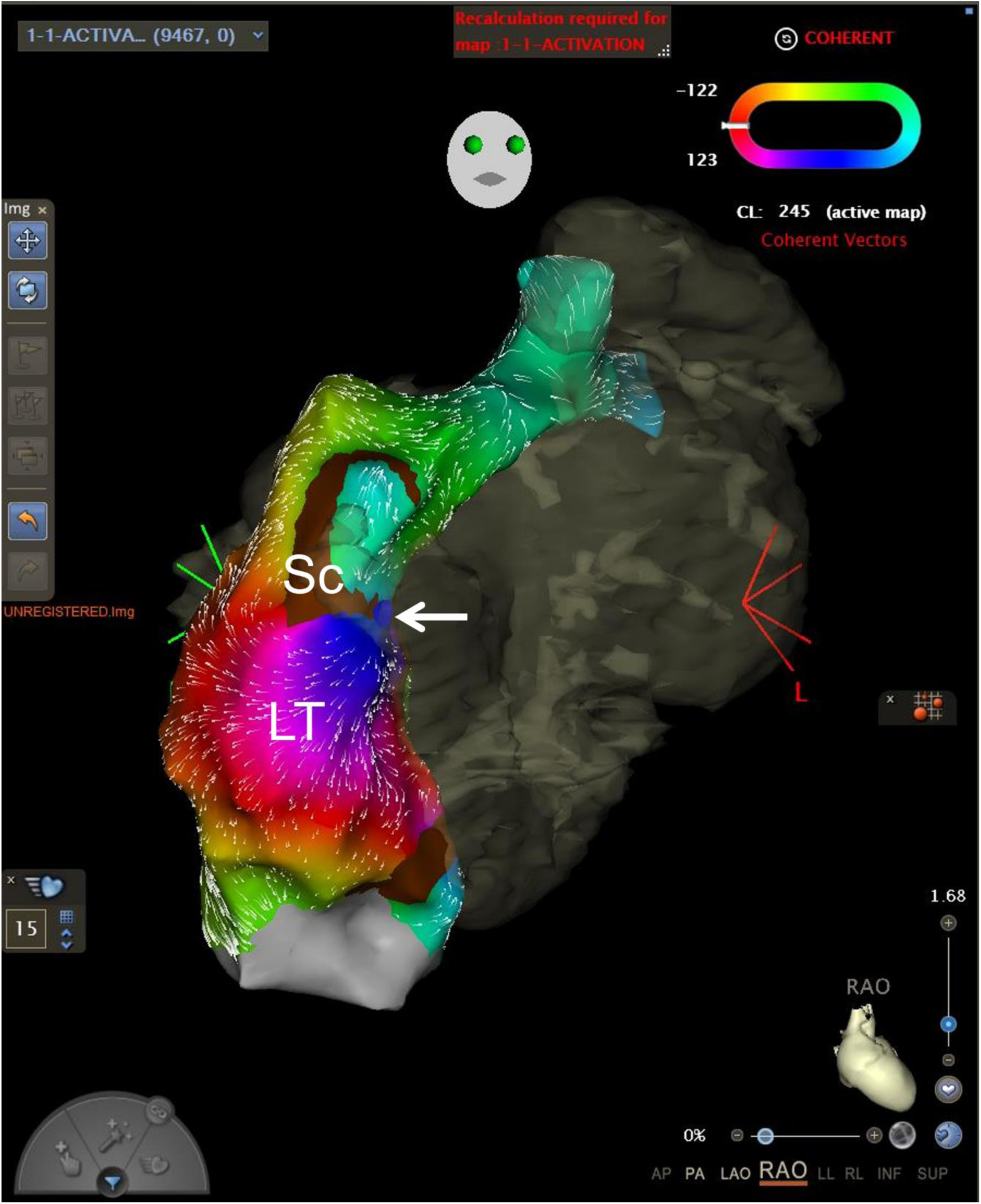
Activation map of RA scar-related reentrant tachycardia in a male in his 20s with LT Fontan for complicated AVSD. The critical isthmus (white arrow) was the suture line for LT formation. AVSD: atrioventricular septal defect; LT: lateral tunnel; RA: right atrium; Sc: scar

**Figure 6.**
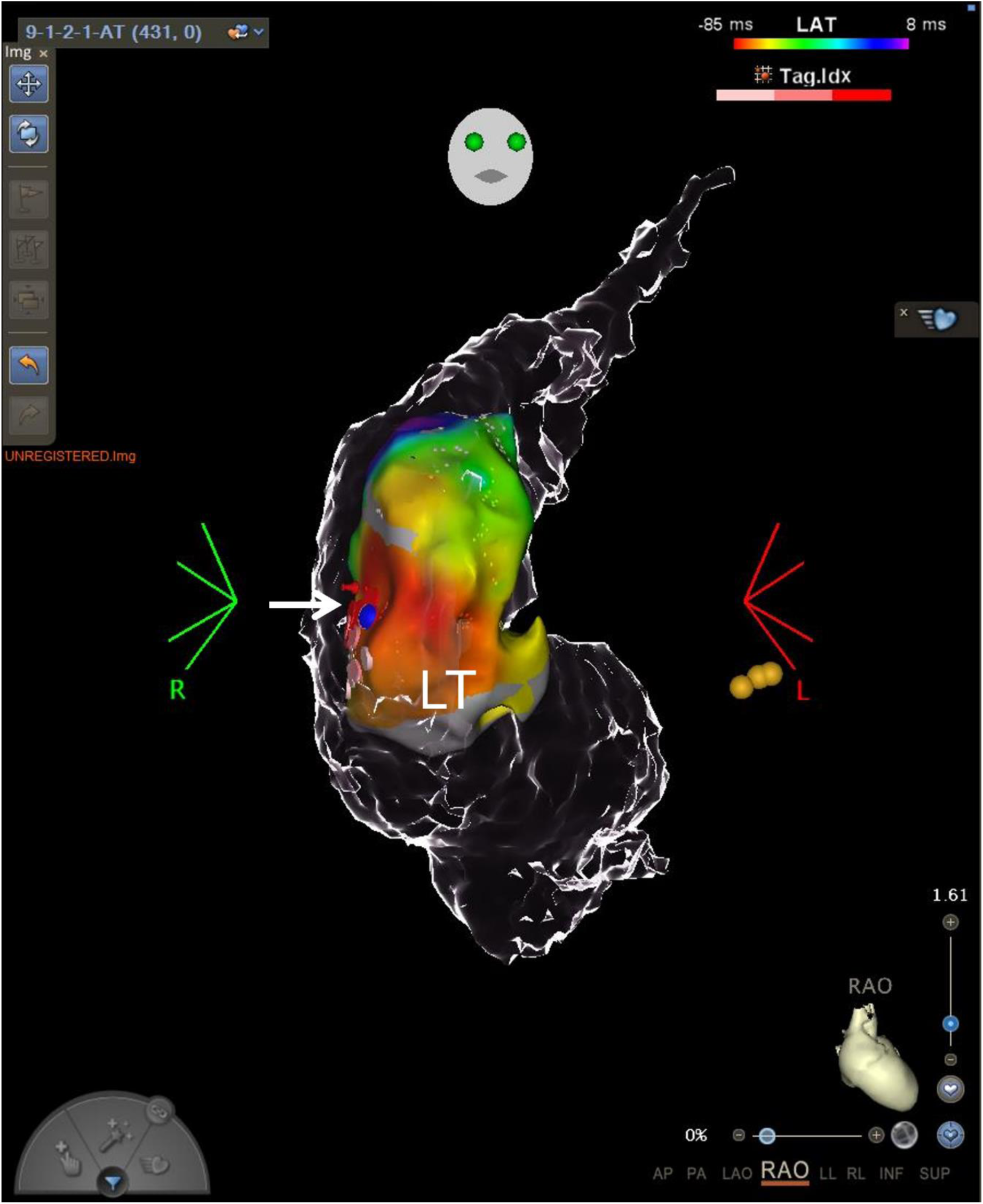
Activation map of focal AT in a male in his 20s with LT Fontan for complicated AVSD. The origin of focal AT (white arrow) was located at the RA lateral wall in the LT. AT: atrial tachycardia; AVSD: atrioventricular septal defect; LT: lateral tunnel; RA: right atrium

**Table 1.** Baseline characteristics of the patients.

|  | Total<br>(n=49) | APC<br>(n=9) | LT<br>(n=19) | ECC<br>(n=21) | *P*<br>value |
| --- | --- | --- | --- | --- | --- |
| Age at procedure (year) | 29.2±10.0 | 38.4±5.6<sub>a</sub> | 28.6±8.4<sub>b</sub> | 25.8±10.6<br>b | 0.004 |
| Male, n (%) | 27 (55.1) | 3 (33.3)<sub>a</sub> | 5 (26.3)<sub>a</sub> | 14 (66.7)<sub>b</sub> | 0.028 |
| Congenital heart disease |  |  |  |  |  |
| Complicated AVSD, n (%) | 12 (24.5) | 1 (11.1) | 5 (26.3) | 6 (28.6) | —<sup>*</sup> |
| Complicated DORV, n (%) | 9 (18.4) | 4 (44.4) | 2 (10.5) | 3 (14.3) | —<sup>*</sup> |
| TA, n (%) | 8 (16.3) | 3 (33.3) | 2 (10.5) | 3 (14.3) | —<sup>*</sup> |
| PA IVS, n (%) | 8 (16.3) | 0 (0.0) | 3 (15.8) | 5 (23.8) | —<sup>*</sup> |
| MA, n (%) | 7 (14.3) | 1 (11.1) | 3 (15.8) | 3 (14.3) | —<sup>*</sup> |
| HLHS, n (%) | 1 (2.0) | 0 (0.0) | 1 (5.3) | 0 (0.0) | —<sup>*</sup> |
| DILV, n (%) | 1 (2.0) | 0 (0.0) | 1 (5.3) | 0 (0.0) | —<sup>*</sup> |
| Ebstein anomaly, n (%) | 1 (2.0) | 0 (0.0) | 1 (5.3) | 0 (0.0) | —<sup>*</sup> |
| Complicated ccTGA, n<br>(%) | 1 (2.0) | 0 (0.0) | 1 (5.3) | 0 (0.0) | —<sup>*</sup> |
| Swiss-cheese VSD, n (%) | 1 (2.0) | 0 (0.0) | 0 (0.0) | 1 (4.8) | —<sup>*</sup> |
| Right isomerism, n (%) | 11 (22.4) | 1 (11.1) | 6 (31.6) | 4 (19.0) | —<sup>*</sup> |
| Left isomerism, n (%) | 2 (4.1) | 0 (0.0) | 1 (5.3) | 1 (4.8) | —<sup>*</sup> |
| Situs inversus, n (%) | 5 (10.2) | 0 (0.0) | 2 (10.5) | 3 (14.3) | —<sup>*</sup> |
| Age at Fontan procedure<br>(year) | 4.6±4.3 | 3.4±3.9 | 3.9±2.9 | 5.7±5.7 | 0.272 |
| Prior Fontan conversion, n (%) | 9 (18.4) | — | 1 (5.3) | 8 (38.1) | —<sup>*</sup> |
| Sinus node dysfunction, n (%) | 13 (26.5) | 6 (66.7)<sub>a</sub> | 3 (15.8)<sub>b</sub> | 4 (19.0)<sub>b</sub> | 0.010 |
| Liver cirrhosis, n (%) | 29 (59.2) | 8 (88.9)<sub>a</sub> | 13 (68.4)<sub>a</sub> | 8 (38.1)<sub>b</sub> | 0.020 |
| Systemic ventricular EF (%) | 52.3±10.9 | 52.0±6.8 | 51.4±12.9 | 53.2±10.8 | 0.887 |
| Time interval from Fontan procedure to ablation (years) | 21.9±10.1 | 27.2±7.7 | 22.1±8.5 | 19.5±11.7 | 0.160 |

APC: atriopulmonary connection; AVSD: atrioventricular septal defect; ccTGA: congenitally corrected transposition of the great arteries; DILV: double-inlet left ventricle; DORV: double-outlet right ventricle; ECC: extracardiac conduit; EF: ejection fraction; HLHS: hypoplastic left heart syndrome; LT: lateral tunnel; MA: mitral atresia; PA IVS: pulmonary atresia with intact ventricular septum; TA: tricuspid atresia; VSD: ventricular septal defect
<sup>\*</sup>Statistical analysis was not conducted owing to the low frequency values.
<sub>a, b</sub> The same subscript letters indicate non-significant differences, and different subscript letters indicate significant differences across the groups based on multiple comparison tests.

**Table 2.** Fontan conduit puncture methods and electrophysiological features.

|  | Total<br>(n=49) | APC<br>(n=9) | LT<br>(n=19) | ECC<br>(n=21) | *P*<br>value |
| --- | --- | --- | --- | --- | --- |
| Fontan puncture method |  |  |  |  |  |
| No need, n (%) | 14 (28.6) | 8 (88.9) | 4 (21.1) | 1 (4.8) | —<sup>*</sup> |
| BRK only, n (%) | 12 (24.5) | 1 (11.1) | 9 (47.4) | 3 (14.3) | —<sup>*</sup> |
| BRK and balloon, n (%) | 5 (10.2) | — | 0 (0.0) | 5 (23.8) | —<sup>*</sup> |
| BRK and cut stylet, n (%) | 4 (8.2) | — | 4 (21.1) | 0 (0.0) | —<sup>*</sup> |
| BRK, cut style, and<br>balloon, n (%) | 13 (26.5) | — | 2 (10.5) | 11 (52.4) | —<sup>*</sup> |
| Transcaval puncture, n (%) | 6 (12.2) | — | 3 (15.8) | 3 (14.3) | —<sup>*</sup> |
| Failed, n (%) | 1 (2.0) | — | 0 (0.0) | 1 (4.8) | —<sup>*</sup> |
| No. of inducible tachycardia | 1.3±0.6 | 1.3±0.5 | 1.4±0.7 | 1.1±0.6 | 0.093 |
| 0, n (%) | 1 (2.0) | 0 (0.0) | 0 (0.0) | 1 (4.8) | —<sup>*</sup> |
| 1, n (%) | 37 (75.5) | 6 (66.7) | 13 (68.4) | 18 (85.7) | —<sup>*</sup> |
| 2, n (%) | 8 (16.3) | 3 (33.3) | 4 (21.1) | 2 (9.5) | —<sup>*</sup> |
| 3, n (%) | 3 (6.1) | 0 (0.0) | 2 (10.5) | 0 (0.0) | —<sup>*</sup> |
| ≥2, n (%) | 11 (22.4) | 3 (33.3) | 6 (31.6) | 2 (9.5) | 0.171 |
| Inducible tachycardia, n (%) | 59 (100) | 10 (100) | 26 (100) | 23 (100) |  |
| IART, n (%) | 23 (39.0) | 8 (80.0)<sub>a</sub> | 10 (38.5)<sub>b</sub> | 5 (21.7)<sub>b</sub> | 0.007 |
| Focal AT, n (%) | 17 (28.8) | 2 (20.0) | 9 (34.6) | 6 (26.1) | 0.641 |
| AVRT, n (%) | 7 (11.9) | 0 (0.0) | 3 (11.5) | 4 (17.4) | —<sup>*</sup> |
| AVNRT, n (%) | 6 (10.2) | 0 (0.0) | 3 (11.5) | 3 (13.0) | —<sup>*</sup> |
| AVRT-TN, n (%) | 6 (10.2) | 0 (0.0) | 1 (3.8) | 5 (21.7) | —<sup>*</sup> |

APC: atriopulmonary connection; AT: atrial tachycardia; AVNRT: atrioventricular nodal reentrant tachycardia; AVRT: atrioventricular reentrant tachycardia; AVRT-TN: atrioventricular reciprocating tachycardia involving twin atrioventricular nodes; BRK: Brockenbrough needle; ECC: extracardiac conduit; IART: intra-atrial reentrant tachycardia; LT: lateral tunnel
<sup>\*</sup>Statistical analysis was not conducted owing to the low frequency values.

**Table 3.** Locations of IARTs and focal ATs.

|  | Total | APC | LT | ECC | *P*<br>value |
| --- | --- | --- | --- | --- | --- |
| IART, n (%) | 23 (100.0) | 8 (100.0) | 10 (100.0) | 5 (100.0) |  |
| RA lateral wall, n (%) | 16 (69.6) | 7 (87.5) | 7 (70.0) | 2 (40.0) | 0.194 |
| CTI, n (%) | 2 (8.7) | 0 (0.0) | 1 (10.0) | 1 (20.0) | —<sup>*</sup> |
| Septum, n (%) | 2 (8.7) | 1 (12.5) | 0 (0.0) | 1 (20.0) | —<sup>*</sup> |
| Coronary sinus, n (%) | 2 (8.7) | 0 (0.0) | 1 (10.0) | 1 (20.0) | —<sup>*</sup> |
| Unknown, n (%) | 1 (4.3) | 0 (0.0) | 1 (10.0) | 0 (0.0) | —<sup>*</sup> |
| Focal AT, n (%) | 17 (100.0) | 2 (100.0) | 9 (100.0) | 6 (100.0) |  |
| RA lateral wall, n (%) | 10 (58.8) | 2 (100.0) | 6 (66.7) | 2 (33.3) | 0.198 |
| Coronary sinus ostium, n (%) | 2 (11.8) | 0 (0.0) | 1 (11.1) | 1 (16.7) | —<sup>*</sup> |
| LA anterior wall, n (%) | 2 (11.8) | 0 (0.0) | 1 (11.1) | 1 (16.7) | —<sup>*</sup> |
| RSPV, n (%) | 1 (5.9) | 0 (0.0) | 1 (11.1) | 0 (0.0) | —<sup>*</sup> |
| Unknown, n (%) | 2 (11.8) | 0 (0.0) | 0 (0.0) | 2 (33.3) | —<sup>*</sup> |
| RA lateral wall as an origin of IART or focal AT, n (%) | 26 (65.0) | 9 (90.0)<sub>a</sub> | 13 (68.4)<sub>a</sub> | 4 (36.4)<sub>b</sub> | 0.033 |

APC: atriopulmonary connection; AT: atrial tachycardia; CTI: cavotricuspid isthmus; ECC: extracardiac conduit; IART: intra-atrial reentrant tachycardia; LA: left atrium; LT: lateral tunnel; RA: right atrium; RSPV: right superior pulmonary vein
<sup>\*</sup>Statistical analysis was not conducted owing to the low frequency values.
<sub>a, b</sub> The same subscript letters indicate non-significant differences, and different subscript
letters indicate significant differences across the groups based on multiple comparison tests.

Table 4 presents the procedural and clinical outcomes of patients. The acute success rate of repetitive procedures and procedure-related complication rate were 73.5% and 4.1%, respectively. Complications included a femoral arteriovenous fistula and minor stroke without long-term sequelae. The acute success rate was significantly higher in the patients who underwent the LT or ECC Fontan procedure than in those who the APC Fontan procedure (*P*=0.010). The recurrence rate was 34.7% over a follow-up period of 78.0±71.9 months. Fontan conversion (APC to ECC) with maze procedure was performed in five patients with the markedly enlarged RA and had IART recurrence. IART recurred after surgery in one of the five patients. Fontan conversion (LT to ECC) with maze procedure was performed in one patient who recurrent IART and atrial fibrillation, with no IART and atrial fibrillation recurrence for 75.8 months post-surgery. The cumulative SVT recurrence rate after repetitive catheter ablations was significantly lower in patients who the LT or ECC Fontan procedure than in those who the APC Fontan procedure (*P*<0.001; Figure 7). Repetitive catheter ablations were required in 28.6% of the total patients. The number of catheter ablations did not significantly differ across Fontan types. Table 5 presents the acute success and recurrence rates for the SVT subtypes. No significant differences were observed in the acute procedure success rates of IART, focal AT, AVNRT, AVRT-TN, and AVRT.

**Figure 7.**
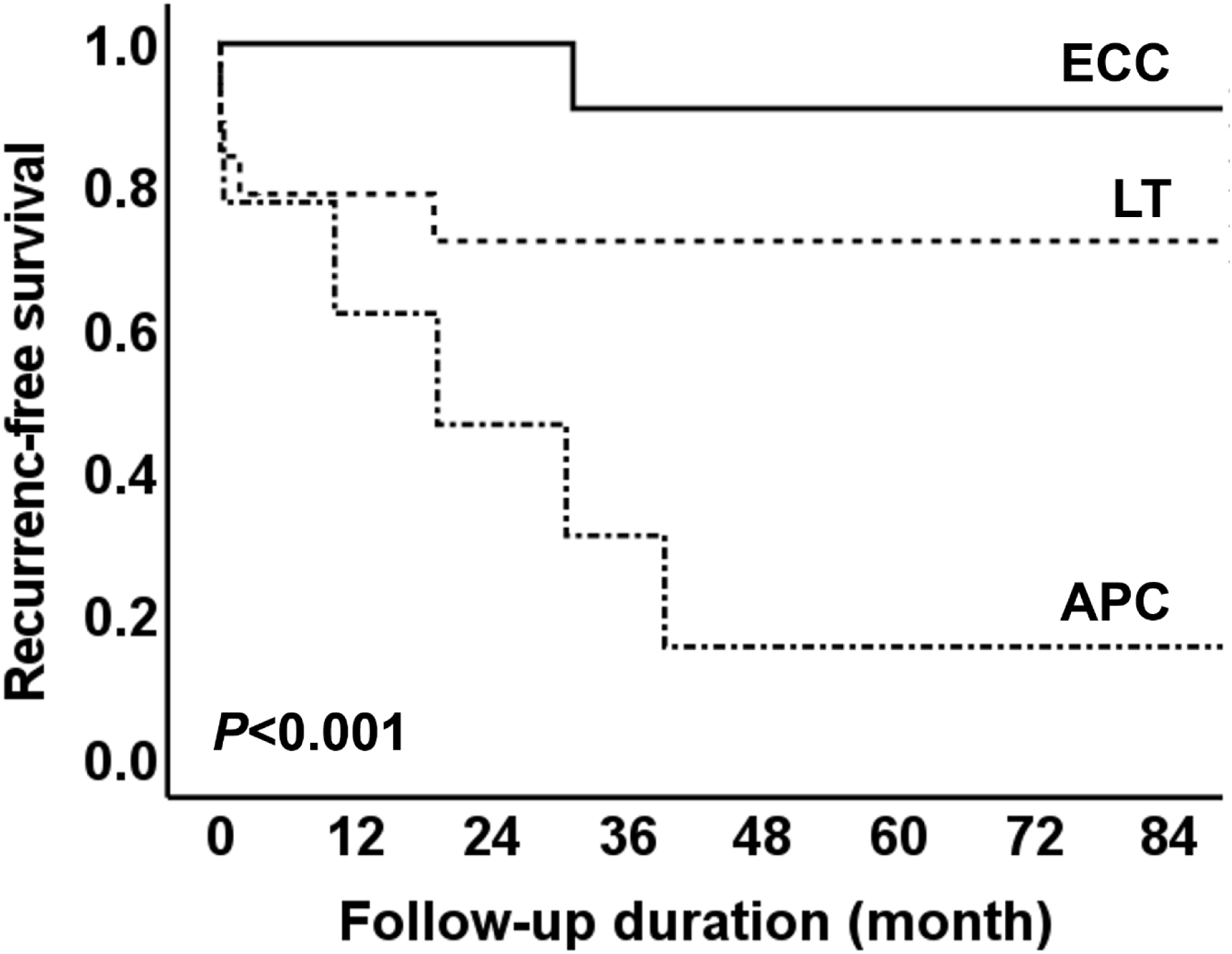
Cumulative SVT recurrence-free survival across Fontan types. APC: atriopulmonary connection; ECC: extracardiac conduit; LT: lateral tunnel; SVT: supraventricular tachycardia

**Table 4.** Procedural and clinical outcomes.

|  | Total<br>(n=49) | APC<br>(n=9) | LT<br>(n=19) | ECC<br>(n=21) | *P*<br>value |
| --- | --- | --- | --- | --- | --- |
| Acute success of repetitive ablation, n (%) | 36 (73.5) | 3 (33.3)<sub>a</sub> | 16 (84.2)<sub>b</sub> | 17 (81.0)<sub>b</sub> | 0.010 |
| Complications, n (%) | 2 (4.1) | 0 (0.0) | 1 (5.3) | 1 (4.8) | —<sup>*</sup> |
| Femoral arteriovenous fistula, n (%) | 1 (2.0) | 0 (0.0) | 1 (5.3) | 0 (0.0) | —<sup>*</sup> |
| Minor stroke, n (%) | 1 (2.0) | 0 (0.0) | 0 (0.0) | 1 (4.8) | —<sup>*</sup> |
| No. of catheter ablation | 1.4±0.7 | 1.3±0.5 | 1.5±0.8 | 1.3±0.8 | 0.238 |
| 1, n (%) | 35 (71.4) | 6 (66.7) | 12 (63.2) | 17 (81.0) | —<sup>*</sup> |
| 2, n (%) | 9 (18.4) | 3 (33.3) | 4 (21.1) | 2 (9.5) | —<sup>*</sup> |
| 3, n (%) | 4 (8.2) | 0 (0.0) | 3 (15.8) | 1 (4.8) | —<sup>*</sup> |
| 4, n (%) | 1 (2.0) | 0 (0.0) | 0 (0.0) | 1 (4.8) | —<sup>*</sup> |
| ≥2, n (%) | 14 (28.6) | 3 (33.3) | 7 (36.8) | 4 (19.0) | 0.434 |
| Recurrence, n (%) | 17 (34.7) | 7 (77.8)<sub>a</sub> | 5 (26.3)<sub>b</sub> | 5 (23.8)<sub>b</sub> | 0.011 |
| Treatment after recurrence, n (%) | 17 (100) | 7 (100) | 5 (100) | 5 (100) |  |
| Medical therapy without further procedure, n (%) | 11 (64.7) | 2 (28.6) | 4 (80.0) | 5 (100.0) | —<sup>*</sup> |
| Fontan conversion with maze, n (%) | 6 (35.3) | 5 (71.4) | 1 (20.0) | 0 (0.0) | —<sup>*</sup> |
| Follow-up period (month) | 78.0±71.9 | 111.0±85. | 79.5±83.5 | 62.4±49.6 | 0.238 |
|  |  | 3 |  |  |  |

APC: atriopulmonary connection; ECC: extracardiac conduit; LT: lateral tunnel \*Statistical analysis was not conducted owing to the low frequency values. <sub>a, b</sub> The same subscript letters indicate non-significant differences, and different subscript letters indicate significant differences across the groups based on multiple comparison tests.

**Table 5.** Procedural and clinical outcomes for the SVT subtypes.

|  | IART<br>(n=23) | Focal AT<br>(n=17) | AVRT<br>(n=7) | AVNRT<br>(n=6) | AVRT-TN<br>(n=6) | *P*<br>value |
| --- | --- | --- | --- | --- | --- | --- |
| Acute success, n (%) | 16 (69.6) | 12 (70.6) | 6 (85.7) | 4 (66.7) | 5 (83.3) | 0.612 |
| Recurrence, n (%) | 8 (34.8) | 6 (35.3) | 3 (42.9) | 2 (33.3) | 1 (16.7) | 0.616 |

AT: atrial tachycardia; AVNRT: atrioventricular nodal reentrant tachycardia; AVRT: atrioventricular reentrant tachycardia; AVRT-TN: atrioventricular reciprocating tachycardia involving twin atrioventricular nodes; IART: intra-atrial reentrant tachycardia; SVT: supraventricular tachycardia

## Discussion

The main findings of the present study are as follows: (1) IART and focal AT were the most common SVTs in patients with Fontan circulation. (2) The RA lateral wall was the most common site of critical IART isthmus and focal AT origin in patients with Fontan circulation. (3) LT baffle and ECC puncture were successfully performed using a Brockenbrough needle, with or without a cut stylet and staged balloon dilation. (4) Catheter ablation for SVT was effective and safe in patients who underwent the LT and ECC Fontan procedures.

### Complex Cardiac Anatomy in Patients with FSV

Patient with FSV usually have various complex congenital heart diseases and conduction system abnormalities, such as atrial isomerism or twin AV nodes.^8–10^ Operators should fully understand the heart anatomy and surgical history before the procedure. Pre-procedural echocardiography, CT, and intraprocedural intracardiac or transesophageal echocardiography are crucial. Access routes and catheter positioning should be planned before the procedure. Similarly, the anatomic relationship between the Fontan pathway and the heart should be noted. The anatomy of a conduction system can differ from that of a normal system. Isomerism should be checked to predict anomalies in the cardiac conduction and venous system. Patients with right isomerism have twin AV nodes that may be potential substrates for AV reciprocating tachycardia.^11–13^

### Arrhythmogenicity in Patients with Fontan Circulation

IART and focal AT are reportedly common in patients with FSV.^14–16^ The evolution of the Fontan procedure from APC to LT and ECC has reduced the incidence of atrial arrhythmia.^4,5^ Patients who have undergone the APC Fontan procedure have a significantly dilated RA because of prolonged exposure to elevated systemic venous pressure. In addition, prior RA incision and sinus node dysfunction contribute to substantial structural and electrical remodeling, creating a highly arrhythmogenic substrate. Catheter ablation for IART and AT is challenging in these patients owing to significant structural and electrical remodeling.^17^ Fontan conversion (from APC to ECC) with maze procedure reduces RA arrhythmogenicity; however, it may persist. The LT Fontan procedure reduces structural and electrical atrial remodeling compared with the APC Fontan procedure; however, this remodeling persists. The structural and electrical atrial remodeling in APC and LT Fontan procedures can cause IART and focal AT to originate from the RA. In contrast, in ECC Fontan, the atria are not exposed to high pressure, and electrical remodeling is significantly reduced. The ECC Fontan procedure was associated with a substantially lower risk of late atrial arrhythmia than the LT Fontan procedure in a large multicenter registry.^18^ The APC Fontan procedure was less common than the LT and ECC Fontan procedures in the study population, possibly because the ECC Fontan procedure was performed earlier in Korea than in other countries. Fontan conversion with maze procedure may be a good treatment option for uncontrolled atrial tachyarrhythmias in patients who have undergone the APC Fontan procedure.^16,19,20^ However, catheter ablation for IARTs or focal ATs could be challenging in patients who underwent Fontan conversion with maze procedure as arrhythmia substrates could be complicated.

### Heart Accessibility in Patients with Fontan Circulation

A considerable number of IARTs or focal ATs originate from the Fontan conduit in patients who underwent the LT Fontan procedure, necessitating a thorough tachycardia mapping of the Fontan conduit before LT Fontan puncture.^21^ The complexity of Fontan conduit puncture is dependent on the underlying heart anatomy and Fontan pathway materials. Puncture of the LT Fontan baffle consisting of the autologous or bovine pericardium is not difficult; however, puncture of the ECC made of expanded polytetrafluoroethylene is challenging. Consequently, the cut stylet and staged balloon dilation are usually required for Fontan puncture (Figure 2).^7^ The Brockenbrough needle stylet is modified through oblique cutting (1–2 mm from the tip), to facilitate puncture of the expanded polytetrafluoroethylene Fontan conduit.^6^ In addition, the stylet is shortened to prevent fracture during puncture. The targeted site and Brockenbrough needle trajectory for Fontan puncture should be determined using heart CT before the procedure. The sternal wire is a useful landmark for determining the puncture level. Intracardiac or esophageal echocardiographic guidance is essential during Fontan conduit puncture. At three-dimensional electroanatomical mapping system and radiofrequency needle can be helpful.^22^ Transcaval cardiac puncture can also be a useful approach for ECC puncture.^23^ For this approach, the anatomical relationship should be confirmed by heart CT.

### Mapping and Ablation in Patients with Fontan Circulation

Catheter placement is limited in patients who have undergone the LT or ECC Fontan procedure owing to the single puncture access. Additionally, reference catheter placement is crucial. In most cases, a stable reference catheter with a stable atrial signal can be achieved by placing the catheter in the left pulmonary artery adjacent to the atrium. Detailed mapping of the Fontan pathway and atria using a three-dimensional electroanatomical mapping system is necessary. Furthermore, His bundle mapping is critical for AVNRT and AVRT-TN catheter ablation.

### Implications of the Study Findings

This study advances our current knowledge on catheter ablation in patients with Fontan circulation, suggesting that careful patient selection and detailed mapping could improve outcomes in this complex population. The findings support the consideration of catheter ablation as a first-line treatment for SVTs in patients who have undergone the LT and ECC Fontan procedures, potentially reducing morbidity and improving quality of life.

### Study Limitations

The detailed procedures varied among the operators owing to the multicenter and retrospective design of the study. However, the main workflow procedures did not differ. The number of patients enrolled in this study was relatively small, and the underlying congenital heart diseases were heterogeneous. These limitations are difficult to avoid because these procedures are not commonly performed in patients with complex congenital heart disease. Further studies are needed to confirm the efficacy and safety of catheter ablation in these patients.

## Conclusions

IART and focal AT originating from the RA lateral wall are common SVTs in patients with Fontan circulation. Furthermore, catheter ablation for SVT is effective and safe in patients who have undergone LT and ECC Fontan procedures.

## Data Availability

The data that support the findings of this study are available from the corresponding author, Jae-Sun Uhm, upon reasonable request.

## Acknowledgments

None

## Sources of Funding

This research did not receive any specific grants from funding agencies in the public, commercial, or not-for-profit sectors.

## Disclosures

There are no conflicts of interest to declare.

## Non-standard Abbreviations and Acronyms

APC: atriopulmonary connection
AT: atrial tachycardia
AVNRT: atrioventricular nodal reentrant tachycardia
AVRT: atrioventricular reentrant tachycardia
AVRT-TN: atrioventricular reciprocating tachycardia involving twin AV nodes
CT: computed tomography
ECC: extracardiac conduit
ECG: electrocardiogram
FSV: functional single ventricle
IART: intra-atrial reentrant tachycardia
LT: lateral tunnel
RA: right atrium
SVT: supraventricular tachyarrhythmia

## Notes

### Competing Interest Statement

The authors have declared no competing interest.

### Author Declarations

This retrospective multicenter cohort study was approved by the Institutional Review Board of Severance Hospital, Seoul National University Children's Hospital, and Sejong General Hospital (IRB number: 4-2025-1595, 2602-118-1718, 2026-02-006).

